# Identifying subtypes of a stigmatized medical condition

**DOI:** 10.1101/19005223

**Authors:** Irene S. Gabashvili

## Abstract

**Background:** Some conditions - such as obesity, depression and functional odor disorders - come with a social stigma. Understanding the etiology of these conditions helps to avoid stereotypes and find remedies. One of the major obstacles facing researchers, especially for those studying socially distressing metabolic malodor, is the difficulty in assembling biologically homogenous study cohorts.

**Objective:** The aim of this study was to examine phenotypic variance, self-reported data and laboratory tests for the purpose of identifying clinically relevant and etiologically meaningful subtypes of idiopathic body odor and the “People are Allergic To Me” (PATM) syndrome.

**Methods:** Participants with undiagnosed body odor conditions enrolled to participate in this research study initiated by a healthcare charity MEBO Research and sponsored by Wishart Research group at the Metabolomics Innovation Centre, University of Alberta, Canada. Primary outcomes were differences in metabolite concentrations measured in urine, blood and breath of test and control groups. Principal component analyses and other statistical tests were carried out for these measurements.

**Results:** While neither of existing laboratory tests could reliably predict chronic malodor symptoms, several measurements distinguished phenotypes at a significance level less than 5%. Types of malodor can be differentiated by self-reported consumption of (or sensitivity to) added sugars (p<0.01), blood alcohols after glucose challenge (especially ethanol: p<0.0005), urinary excretion of phenylalanine, putrescine, and combinations of blood or urine metabolites.

**Conclusions:** Our preliminary results suggest that malodor heterogeneity can be addressed by analyses of phenotypes based on patients’ dietary and olfactory observations. Our studies highlight the need for more trials. Future research focused on comprehensive metabolomics and microbiome sequencing will play an important role in the diagnosis and treatment of malodor.

**Trial Registration:** The study discussed in the manuscript was registered as NCT02692495 at clinicaltrials.gov. The results were compared with our earlier study registered as NCT02683876.

## Introduction

Certain body and breath odors are known to indicate serious infections and life-threatening conditions. Malodors without accompanying physical symptoms, however, could be a sign of psychologically but not physically debilitating errors of metabolism. One example is Trimethylaminuria (TMAU) leading to excessive excretion of foul-smelling trimethylamine (TMA) in the sweat and breath. Even this relatively straightforward disorder of choline metabolism exhibits complex genetic and environmental regulation. It is no longer thought to be exclusively caused by mutations in the gene coding for liver enzyme FMO3 (flavin-containing monooxygenase isoform 3), as other genes such as oxidoreductases [1] and environmental exposures may be also implicated.

TMAU patients could be physically healthier than the average population, unlike patients of other diseases with altered levels of TMA and TMAO (Trimethylamine-N-Oxide) such as Chronic Kidney Disease (CKD) and acute coronary syndromes. One might even expect that sufferers of TMA-odor develop gut microbiota that potentially protects them from cardiovascular disease [2]. The same could apply to other sufferers of idiopathic malodor – for example, children with bad breath but good teeth [3] or underarm odor sufferers that harbor less acne bacteria [4]. There is no indication that halitosis caused by a mutation in a cancer-protecting gene SELENBP1 increases risk of cancer [5]. Yet, psychological, social, and economic implications of uncontrollable odors can have a devastating impact on health [6].

The rise of online health communities enabled sufferers of rare diseases to communicate with their peers around the world and exchange first-hand knowledge of their conditions. Social media is now a key tool educating on unexplained personal malodor production and PATM conditions and could be used as the next disruptive force in clinical trials [7]. We need better diagnostic tests, as the most readily available diagnostic test, measurement of urinary TMA levels after a choline challenge, is only applicable to one condition – trimethylaminuria - and is useful for less than 30% of sufferers with the most severe TMAU-like cases [8; Nigel Manning, Sheffield Children’s Hospital and Maria de la Torre, MEBO Research, personal communication].

Not having a diagnosis takes an enormous emotional toll. To engage researchers in projects addressing community needs, healthcare charity MEBO Research identified non-standard diagnostic tests that typically yielded positive results for sufferers of socially-disabling malodor, and initiated several studies. In this paper, we present the most common patterns observed.

## Methods

### Recruitment

Over 300 volunteers from all racial and ethnic groups expressed interest in Internet-based medical research of malodor related conditions and participated in our online surveys. Many of the participants shared results of laboratory tests they had taken in hopes to find the primary cause of their malodor. Internationally-recruited volunteers with undiagnosed body odor and halitosis enrolled to participate in several diagnostic studies, some of which are registered at clinicaltrials.gov. Participants underwent various blood, breath and urine tests.

57 Canadian residents expressed interest in the exploratory metabolomic testing of urine for the study registered as Clinical Trials NCT02683876. The study was reviewed by MeBO Research IRB00010169 (FWA00026547 expires on 02/14/2023), registered with The Food and Drug Administration (FDA) and Office for Human Research Protections (OHRP) through a system maintained by the US Department of Health and Human Services (DHHS), as required under 21 CFR part 56, subpart B, and 45 CFR part 46, subpart E. (Clinical Protocol Number: 201502030014MEBO). All necessary participant consent has been obtained and the appropriate institutional forms have been archived. Wishart Laboratory at the Metabolomics Innovation Centre shipped kits to 27 enrollees that re-confirmed their interest and availability. 17 participants returned the kits with their urine samples, 15 of which were considered viable. The samples were of 10 women and 5 men, aged 22-55 years, who experienced symptoms for 4-30 years. The first appearance of malodor symptoms was about 5 years earlier in male than female participants. The mean duration of symptoms was 12 years.

### Analysis

Our previous exploratory testing included Gut Permeability Profile, Gut Fermentation Profile, Hydrogen Breath test, functional blood B vitamin profile, plasma D-lactate, and urine Indicans [10, 11]. For this study, Wishart laboratory performed DI/LC-MS/MS analysis of morning urine samples [12, 13]. By using their assay, a total of 96 metabolites of different classes (amino acids, biogenic amines, acylcarnitines, glycerophospholipids, sphingolipids) were identified and quantified. The results were normalized with creatinine concentrations and compared to the results of 22 healthy volunteers. MeBO Research IRB served as an independent ethics committee to protect the privacy and confidentiality of data concerning the research subjects. A set of R packages and Excel macros was developed for statistical analysis. Aurametrix software was used to analyze dietary patterns.

## Results and Discussion

### Odor biomarkers

We identified 95 metabolites in 15 samples, including Amino Acids, Biogenic Amines, Acylcarnitines, Glycerophospholipids and Sphingolipids. The ranges of these metabolites in urine of MEBO sufferers were not statistically different from the ranges in healthy volunteers, although the averages and deviations were different for some compounds.

Participants of our earlier study NCT02692495 underwent a variety of biochemical tests quantifying about two dozen odorous metabolites, by-products of microbial metabolism, in blood and urine. The molecules studied include D-lactate, Indoxyl sulfate (Indican), primary, secondary, and tertiary alcohols, short chain fatty acids and related substances.

We summarized these laboratory test results in a 16×14 table with participants numbered from 1 to 16 and 14 metabolites that were identified in participant samples outside expected ranges.

We replaced actual measured values of metabolites with discrete “buckets” ranging from “-1” (when test results were significantly lower than in healthy participants not complaining of odors) to 1 (values significantly higher than normal).

We analyzed results of all our laboratory test results with several statistical techniques known to bring out strong patterns in a dataset. Results of one such procedure, a principal component analysis (PCA), are shown in Fig.1.

Our main finding was that PCA clustered subjects in groups similar by malodor symptoms reported by the subjects, regardless of the source, age, gender, or other medical history. Another remarkable difference between the two groups was in the participants’ average consumption of sugar. We will refer to the two main clusters identified as the “sweet” group (darker circles clustered near the top of the cube, in Fig. 1, representing participants with higher sugar intake) and the “sour” group (lighter circles near the bottom, with less sugar in their diets). As actual odors reported by the participants of study NCT02692495 featured over a dozen of keywords, we associated the symptoms with a gamut of gray colors ranging from the lightest – starting from “garbage” odor (lightest circles), followed by “fishy”, “ammonia”, sour acetone (body odors associated with alcoholism), and fecal-diarrheal, followed by generic “fecal odor” - the darkest three circles in the “sour” cluster. The “sweet” group symptom keywords mostly include sweetish sulfurous smells. The odors, from the lightest to the darkest, are “sulfur”, “rotten vegetables”, “rotten eggs”, cheesy/sweaty-fecal, sewage, smoke-like, gasoline, “burning”, followed by those who could not smell themselves and, finally, the “PATM” condition shown as black circles (odors could not be detected or named, but people near the sufferer exhibited increased displeasure, coughing, sneezing, and rubbing their noses). We note that low level exposures to sulfur compounds are known to cause similar issues associated with mild dyspnea.

**Figure 1.**
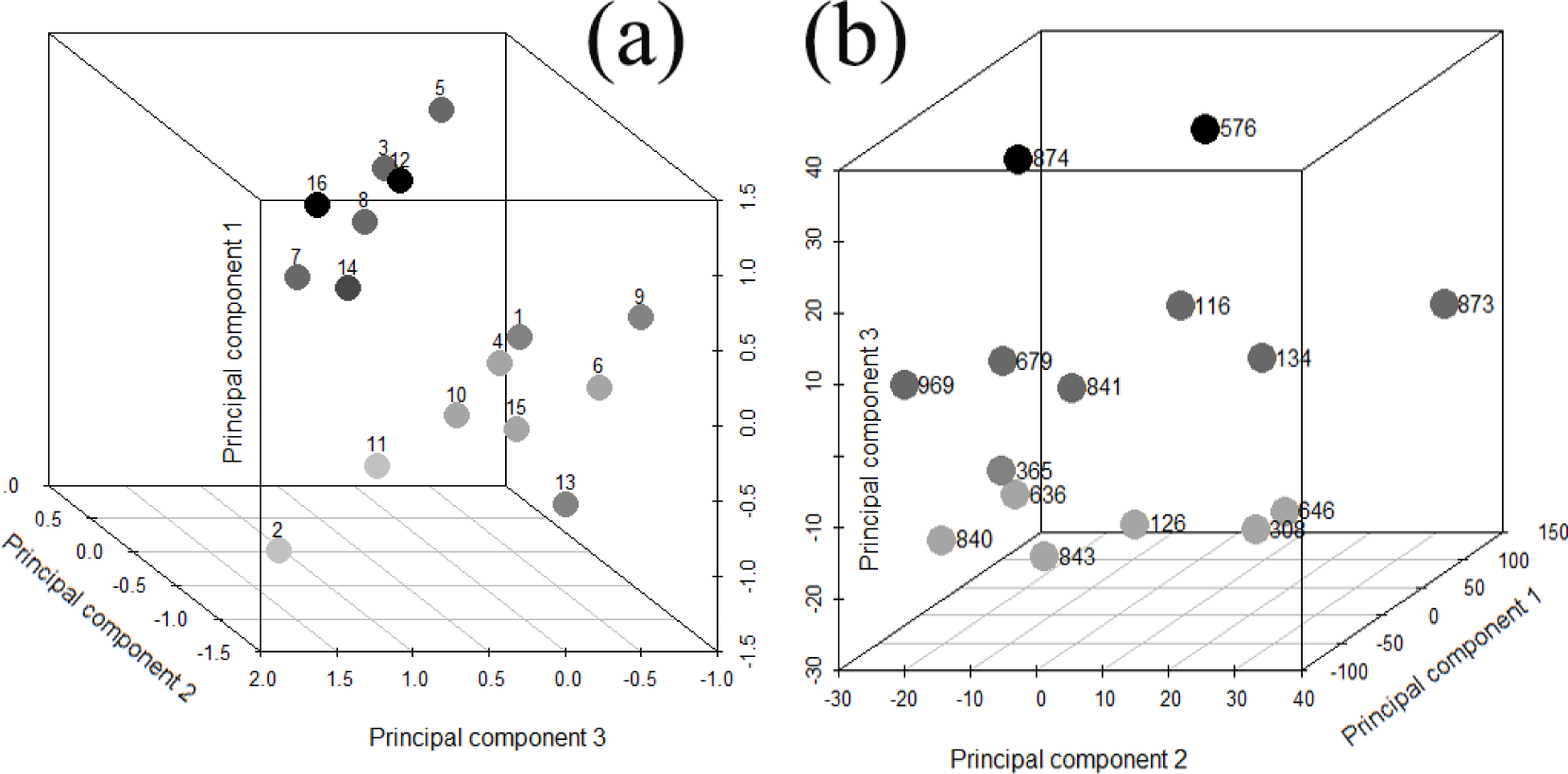
Principal components analysis plot in 3D. The axes show the first three principal components. Each principal component is a linear combination of the original test results indicating over- or under-expression of 19 metabolites measured in blood and urine (a, NCT02692495) or values of 96 urine metabolites normalized to creatinine concentrations (b, NCT02683876). Each dot represents a research participant colored per odors reported (see text for more details).

The axes in Fig. 2a - principal components 1, 2 and 3 - capture a little over 50% of variability in the dataset. Principal component 1 points from sour and acidic odors in detected metabolites (main negative contributors are sour-whiskey-smelling 2-methyl-1-butanol, buttery 2,3-butylene glycol, window-cleaner-like 2-butanol and plastic 2-methyl-1-propanol) towards mild alcohol-like smells such as of 1-propanol. Principal component 3 is somewhat similar – but while its negative values assume more acidic, vinegary, and sour whiskey notes, larger positive values align with other types of sour smells, with sweetish and alcoholic notes. The “sour” subgroup of study participants occupies acidic and sour corner of principal components of the metabolite space, stemming from the lowest values of both PC1 and PC3.

Values of principal component 2 increase as the rancidity and staleness of odors increase – with large positive contributions from rancid-butter-smelling metabolite Butyrate, cheesy Propionate, pungent Acetate and fecal Indican. The “sweet” subgroup is clustered around the areas of highest PC1 (sweet and alcohol-like smells) and PC2.

Another noteworthy observation is that the “sweet” and “sour” groups were significantly different in regards to their sensitivity to probiotic supplements. Three participants from the “sweet” group reported that probiotics improved their odor and only one participant said that the odor worsened after probiotics and vitamins where added to a low choline diet. Neither participant from the “sour” group associated probiotics with improved odor, one participant reported odor getting worse with probiotics but better with plain Greek yogurt, and one participant reported worse odor with antibiotics. Observations about the relationship between odor and meat, dairy, complex and simple carbohydrates or alcohol consumption did not differ between the two groups.

Body levels of B2 vitamin (EGR activation)) were also somewhat correlated with the type of odor. Most deficiencies were observed in the “sweet” group. Borderline riboflavin deficiency has been also known to cause acute lactic acidosis with symptoms such as sweet-smelling breath.

We note that most existing diagnostic tests on their own were poor separators of self-reported odors and, possibly, its underlying causes. Overexpression of Indican, one of the compounds responsible for the foul smell of feces somewhat correlated with complaints of “fecal-sulfuric” vs other types of fecal odors. Still, the keyword “fecal” was the least descriptive in our study. Plasma D-lactate present at over 150 umol/L could be a good predictor of “garbage” odor, but we don’t have enough data to make definite conclusions. Short bowel syndrome, one of medical conditions associated with overproduction of D-lactic acid, does contribute to unpleasant odors with sour notes, described as “fishy”, “feculent” and “pungent”. Lactic acid molecule, itself, smells musky or mousy, like sour milk – contributing to the “sourness” of the odor.

Remarkably, preliminary results of our more recent study NCT02683876 (Fig.2b) were very similar to NCT02692495 and we used the same color palette for odor descriptors in both cases.

The lighter-colored sour group participants (clustered near the bottom of the PCA box in Fig.2b) self-described their odors as mostly vinegary, ammonia, fishy, sour and urine-like odors. When the usual odor was less sour (but nonetheless unpleasant), it became sourer with lower blood sugar (for example, with alpha lipoic acid supplements). Other odor descriptors used in this group were cheese/sweaty, smelly armpit, private-part-related, garbage, musty, metallic, resembling decaying drain pipes, sulfur, rotten egg and skunk.

All members of the second, darker, cluster (positive values of the 3^rd^ principal component, not exceeding 20), mentioned “burning” odors in their self-reports (smoke, cigarette-like, burning, rubbery), even though neither of them was a smoker. Other odor descriptors were animalistic, garbage, mold, sewage, tuna-fishy, smelly socks, feet, rotten eggs, horse stable and fecal.

The “sweet” group had higher sensitivity to sugars (as evident from self-reported dietary observations, in several cases confirmed by hydrogen breath tests) but higher tolerance to dairy than the “sour” group – even though too much dairy could be problematic for both groups. The “sweet” group was also more likely to report heat as a trigger of their odor.

The third group (black circles on the top of the PCA box in Fig.1b) was very different from the rest of the set by their dietary patterns (vegan). The participants appeared to have most food sensitivities and were the only ones diagnosed with non-functional gastrointestinal issues that they had under control.

Notable differences between the groups included lower concentration of putrescine in the “sweet” group (even though all participants had lower levels of this unpleasantly-smelling chemical in their urine than control group). The third group had significantly higher (but still normal) level of excreted Glutamine.

About 20% of participants of the above discussed studies tested positive or borderline positive for secondary TMAU. Metabolic profiling showed that they had significantly higher level of Methionine-Sulfoxide than those who tested negative. This metabolite was in normal range or slightly lower than normal in all study participants.

## Conclusion

Our study provides evidence that community research is an important starting point for exploring elusive disorders but a broader awareness is needed to reduce stigma and conduct in-depth research.

Diet is known to be associated with body odors. A recent study [17] linked greater fruit and vegetable intake to more pleasant sweet- and medicinal-smelling sweat, and, to a lesser extent, so was fat, meat, egg, and tofu intake, while greater carbohydrate intake was making sweat less pleasant. Our participants, however, found that meat, dairy, fruit and vegetables, probiotics, prebiotics, even low-choline and restrictive vegan dieting could make them smell worse, proving that a more individualized dietary approach is needed. Low sugar diets, too, do not reduce odors for everyone, as seen from our study: despite other health benefits, lowering blood sugar didn’t change the unpleasant perception of odors for the “Sour” subgroup of participants, and sometimes even made it worse. This is in line with a recent observation that halitosis sufferers eating less sweets had more unpleasantly-smelling volatile compounds in their breath, while children with caries habitually consuming sugar-containing snacks did not have halitosis [3].

Existing diagnostic tests are not sufficient for diagnosing underlying causes of idiopathic malodor. Intestinal permeability profiling provides marginal value for malodor sufferers, and only helps to differentiate patients with bad breath vs cutaneous respiration. Trimethylaminuria challenge test is currently the highest-yielding diagnostic procedure with less than 30% positives detected [8], but it is not sufficiently reliable, neither is testing for known genetic mutations nor to defined gut microbiome [1, 18]. Glucose challenge test was positive in about half of the participants, but its results could be inferred from a dietary sensitivities questionnaire. Comprehensive profiling of urine metabolites such as Indoxyl sulfate, biogenic amines, lipids and amino acids would help to identify metabolic pathways responsible for malodor. More precise metagenomic sequencing would be also useful.

Our study is the first -omics exploration of systemic body odor suggesting that current expensive, inconvenient and time-consuming challenge tests could be replaced by more efficient and informative assays. Our findings emphasize the importance of patients’ self-reported observations and demonstrate the need for comprehensive metabolome and microbiome investigation toward the successful development of new tests and therapies.

## Data Availability

Data is available from this location:

https://aurametrix.com/NCT2683876.html

## Acknowledgments

The author received no funding for this work and would like to thank the volunteers of the studies described here, Dr. David Wishart’s team at the Metabolomics Innovation Centre (TMIC) for generous support, sample processing and data collection, and staff of MeBO Research, especially Maria de la Torre and Paul Jenkins, for initiating the testing and supporting the study.

